# Topical fresh *Taraxacum mongolicum* wet dressing as an adjunct to ceftriaxone for localized skin and soft tissue infections: A single-center assessor-blinded randomized controlled trial

**DOI:** 10.64898/2026.06.18.26355939

**Authors:** Yi Wang, Xiaoqing Xian, Shengjun Nie, Shengcheng Ma, Han Yang

## Abstract

**Background:** Localized skin and soft tissue infections may need systemic antibacterials, but local inflammation can delay symptom recovery. We evaluated whether topical fresh *Taraxacum mongolicum* wet dressing added to ceftriaxone was associated with short-term benefit in selected clinically stable adults.

**Methods:** In this single-center, assessor-blinded, three-arm randomized trial, 180 adults aged 18-74 years were randomized 1:1:1 to topical *T. mongolicum* plus intravenous ceftriaxone, topical *T. mongolicum* alone, or ceftriaxone alone for 7 days. The primary outcome was day-7 clinical response assessed by blinded independent assessors using prespecified global clinical improvement criteria. Analyses followed the intention-to-treat principle; sensitivity analyses assessed robustness.

**Results:** Day-7 clinical response rates were 91.67% (55/60), 76.67% (46/60), and 68.33% (41/60) in the combined, *T. mongolicum*, and ceftriaxone groups, respectively (overall P = 0.006). Compared with ceftriaxone alone, combined therapy had a higher response rate (risk difference, 23.3 percentage points; 95% CI, 9.6 to 37.0; risk ratio, 1.34; 95% CI, 1.11 to 1.62). Sensitivity analyses were directionally consistent. Secondary outcomes and bacterial clearance favored the combined group. No serious adverse events were reported.

**Conclusions:** In selected clinically stable adults with localized skin and soft tissue infections, adjunctive topical fresh *T. mongolicum* plus ceftriaxone was associated with improved short-term outcomes compared with ceftriaxone alone. Findings require cautious interpretation because this was a single-center, partially blinded trial without a placebo dressing control. The dressing should not replace antibiotics, drainage, or urgent care when indicated.

**Trial registration:** International Traditional Medicine Clinical Trial Registry, ITMCTR2026000549.

## 1. Introduction

Skin and soft tissue infections (SSTIs) are among the most common infectious conditions encountered in clinical practice, encompassing a wide spectrum of diseases ranging from superficial inflammation to more extensive tissue involvement [1,2]. Despite the widespread use of systemic antibiotics and standardized guideline-based management, clinical treatment remains challenging in certain cases, particularly due to persistent local inflammation, delayed symptom resolution, and the increasing prevalence of antimicrobial resistance, including methicillin-resistant *Staphylococcus aureus* (MRSA) [3,4].

In addition to systemic therapy, adjunctive local interventions have been proposed to enhance treatment outcomes by targeting localized inflammatory responses and improving tissue microenvironment. Persistent local edema, impaired microcirculation, and ongoing inflammatory activation may contribute to suboptimal clinical recovery even under appropriate antibiotic treatment [1,2]. However, high-quality randomized controlled evidence supporting the clinical value of adjunctive local therapies in SSTIs remains limited.

*Taraxacum mongolicum* is a traditional medicinal plant widely used for inflammatory conditions in East Asian medicine. Phytochemical and pharmacological studies have identified multiple bioactive constituents, including phenolic acids, flavonoids, and triterpenes, which exhibit antibacterial, anti-inflammatory, antioxidant, and immunomodulatory properties [5–9]. These findings provide a biological rationale for evaluating standardized topical *T. mongolicum* preparations as botanical adjuncts for localized infectious and inflammatory disorders.

Nevertheless, clinical evidence supporting the use of topical *T. mongolicum* in SSTIs remains insufficient, particularly from randomized controlled trials with standardized botanical quality control. Therefore, the present study aimed to evaluate short-term clinical outcomes and safety of topical fresh *T. mongolicum* wet dressing, alone or in combination with systemic ceftriaxone, in selected adults with localized SSTIs. The primary objective was to determine whether adjunctive topical botanical therapy was associated with improved day-7 clinical response compared with ceftriaxone monotherapy. Secondary analyses assessed inflammatory biomarkers, symptom recovery, bacterial clearance, and exploratory heterogeneity in treatment response across anatomical lesion sites.

## 2. Materials and Methods

### 2.1. Trial Registration and Ethical Approval

This randomized controlled trial was registered with the International Traditional Medicine Clinical Trial Registry (ITMCTR2026000549; registration date: 16 January 2026) after participant enrollment had begun and before study completion, data lock, and formal statistical analysis. The delayed public registration resulted from an administrative delay in completing registry submission during trial initiation. The study protocol, eligibility criteria, intervention regimens, primary outcome, planned sample size, rescue-treatment criteria, safety-monitoring procedures, and statistical analysis plan had been finalized before enrollment of the first participant and remained unchanged after registration. All analyses reported in this manuscript were performed after registration. The authors confirm that this is the only clinical trial arising from this protocol and that no related unregistered clinical trial exists.

The study protocol was approved by the Ethics Committee of the 943rd Hospital of the Joint Logistic Support Force of the Chinese People’s Liberation Army (Ethics Approval No.: PLA943EC24002; approval date: 8 November 2024). The ethics review included evaluation of preclinical pharmacological evidence, topical safety data, prior clinical observations, and restriction of topical monotherapy to selected patients without systemic complications or mandatory drainage indications. All enrolled participants were aged 18 years or older. All procedures were conducted in accordance with the Declaration of Helsinki [10]. Written informed consent was obtained from all participants before enrollment. This report follows CONSORT 2025 reporting guidance, and intervention descriptions were prepared with attention to the TIDieR recommendations [11,12].

Patients and the public were not formally involved in the design, conduct, reporting, or dissemination planning of this trial.

### 2.2. Participants

A total of 180 adults with localized skin and soft tissue infections (SSTIs) admitted to our hospital between November 2024 and March 2026 were enrolled. Inclusion criteria were: (1) fulfillment of Western diagnostic criteria for SSTIs per the 2009 Consensus on the Diagnosis and Treatment of Skin and Soft Tissue Infections and 2014 Infectious Diseases Society of America guidelines [1,2], plus traditional Chinese medicine (TCM) diagnostic criteria for “sores and ulcers” with heat-toxin accumulation syndrome based on the Criteria for Diagnosis and Therapeutic Effect of TCM Syndromes [13]; (2) age 18-74 years; (3) lesion swelling area 2-100 cm²; and (4) clinical stability without sepsis, necrotizing infection, deep tissue involvement, hemodynamic instability, or emergency surgical indications. The TCM diagnostic framework was used as an additional eligibility characterization and was not used as evidence of mechanism. For standardized baseline characterization, lesion site, baseline swelling area, and baseline visual analogue scale (VAS) pain score were recorded before randomization. Exclusion criteria were: (1) concurrent sepsis; (2) severe cardiac, hepatic, or renal dysfunction, uncontrolled diabetes mellitus, immunodeficiency, or malignant tumors; (3) pregnancy or lactation; and (4) known allergy to cephalosporins or Asteraceae plants.

### 2.3. Randomization and Blinding

Eligible patients were randomly assigned to the combined group, *T. mongolicum* group, or ceftriaxone group in a 1:1:1 ratio via block randomization (block size = 6; n = 60 per group). Randomization was performed by an independent statistician using a computer-generated sequence. Allocation concealment was ensured using sequentially numbered, opaque, sealed envelopes. Investigators responsible for enrollment opened the next envelope only after eligibility confirmation and written informed consent; they did not have access to future allocations. Randomization was stratified by lesion site and included head/face/neck, trunk, extremities, and perineal/gluteal lesions.

An assessor-blinded design was adopted. Outcome assessors and statisticians were blinded to group allocation, whereas patients and treating physicians were not blinded due to the different routes of administration (topical vs intravenous). To minimize assessment bias, clinical outcome assessments were conducted by blinded independent dermatologists who were not involved in treatment delivery or rescue-treatment decisions and who used prespecified assessment criteria. The three-arm design was used to evaluate the adjunctive value of topical *T. mongolicum* wet dressing when added to ceftriaxone-based management, while also providing supportive information on the independent local therapeutic contribution of topical *T. mongolicum* in a restricted clinically stable SSTI population.

### 2.4. Interventions

All patients received a 7-day treatment course. Patients in the ceftriaxone monotherapy group received intravenous ceftriaxone sodium once daily in accordance with the Guidelines for Clinical Application of Antibacterial Drugs (2015 edition) [14]. The dosage was 2.0 g/day for adults weighing ≥60 kg and 1.5 g/day for adults weighing <60 kg. Ceftriaxone sodium was dissolved in 100 mL of 0.9% sodium chloride solution and administered intravenously. No topical anti-infective intervention was applied in this group.

Patients in the *T. mongolicum* group received fresh *T. mongolicum* wet dressing only, without routine intravenous antibiotics or other topical anti-infective agents. This topical monotherapy arm was restricted to participants who, after guideline-based clinician assessment, had localized and clinically stable infection and did not meet predefined criteria for mandatory systemic antibacterial therapy, urgent incision and drainage, or surgical intervention. Participants with systemic infection, deep tissue infection, necrotizing infection, hemodynamic instability, rapidly progressive disease, or a purulent collection requiring drainage were not eligible for topical monotherapy. This arm was included solely to explore the localized contribution of topical *T. mongolicum* wet dressing in a carefully selected low-risk population under close monitoring with predefined rescue criteria. This design should not be interpreted as recommending antibiotic-free management for SSTIs when systemic antibacterial therapy, drainage, or urgent care is clinically indicated. Fresh *T. mongolicum* paste used for each application was prepared from authenticated fresh plant material that met the predefined batch-level quality-control criteria described in Section 2.5. Fresh *T. mongolicum* paste was wrapped in a single layer of sterile gauze and applied directly to swollen areas in patients without open wounds. For superficial wounds not requiring surgical drainage, petrolatum gauze was used to isolate the wound surface before applying the wet dressing to surrounding intact skin.

Each application lasted 20 minutes. The wet dressing was applied twice daily for all lesion sites, including head/face/neck, trunk, extremities, and perineal/gluteal lesions. Patients in the combined group received both systemic and topical treatments, including intravenous ceftriaxone sodium administered using the same dosage and regimen as in the ceftriaxone group, as well as fresh *T. mongolicum* wet dressing applied in the same manner, duration, and twice-daily schedule as in the *T. mongolicum* group.

A rescue-treatment rule was implemented to preserve patient safety. If fever, progressive erythema, rapidly enlarging swelling, worsening pain, newly formed purulent collection requiring drainage, abnormal vital signs, or laboratory evidence suggesting systemic infection developed during treatment, systemic antibacterial therapy and/or surgical drainage was initiated immediately according to standard care. Such cases were recorded as treatment failure for the purpose of analysis or protocol discontinuation in the intention-to-treat analysis.

### 2.5. Standardization and Quality Control of the *T. mongolicum* Preparation

To ensure consistent quality and traceability of the topical botanical preparation used in the combined and *T. mongolicum* groups throughout the study period, fresh whole *T. mongolicum* plants were obtained in batches from authenticated local sources. The whole plant, including roots, leaves, and flowers, was used for preparation.

Each batch was botanically authenticated before clinical use by the Chief Pharmacist of the Pharmacy Department, 943rd Hospital of the Joint Logistic Support Force of the Chinese PLA, according to the morphological criteria and quality standards described in the 2020 edition of the Pharmacopoeia of the People’s Republic of China [15]. Representative voucher specimens from the initial and subsequent seasonal sources were retained in the specimen room of the Pharmacy Department. The initial representative voucher specimen was assigned the voucher number TM-VS-2024-1105, whereas clinical application batches were recorded separately using batch IDs. Detailed batch-level quality-control records are provided in S3 File.

Fresh plant materials were processed within 2 h after collection or receipt. After washing and draining, the plants were crushed into a granular paste and adjusted with sterile 0.9% sodium chloride solution to maintain appropriate moisture content. The prepared paste was stored at 4 °C before use and was used within 24 h after preparation. Any unused paste exceeding the predefined storage period or showing discoloration, odor change, visible contamination, or abnormal texture was discarded.

Batch records were maintained throughout the trial, including batch number, source type, collection or receipt date, processing date and time, storage duration, clinical use period, botanical authentication result, and quality-control test results. High-performance liquid chromatography (HPLC; Agilent 1260 system; Agilent Technologies, Santa Clara, CA, USA) was used to determine chlorogenic acid content in accordance with the Pharmacopoeia of the People’s Republic of China (2020 edition) [15]. Across all clinically used batches, the mean chlorogenic acid concentration was 5.23 ± 0.44 mg/g. These procedures ensured identity, freshness, traceability, and batch-to-batch consistency of the topical paste used for clinical application.

### 2.6. Outcome Measures

#### 2.6.1. Primary Outcome

The primary outcome was the day-7 clinical response rate, defined as the proportion of participants classified as responders, including complete, major, or partial response. Clinical response categories were assigned by blinded independent assessors using prespecified global clinical improvement criteria adapted from conventional traditional Chinese medicine clinical efficacy evaluation principles [13] and operationalized before trial initiation. The assessment integrated overall improvement in local inflammatory manifestations, including swelling, erythema, local heat, pain, and local functional impairment. Complete response was defined as disappearance or near-disappearance of local inflammatory signs with normalization or near-normalization of local function. Major response was defined as marked improvement in local inflammatory signs and local function. Partial response was defined as clinically meaningful but incomplete improvement in local inflammatory signs. Non-response was defined as insufficient improvement, clinical worsening, discontinuation for clinical deterioration, or requirement for rescue treatment during the study period.

#### 2.6.2. Secondary Outcomes

Secondary outcomes included percentage reduction in lesion swelling area, change in pain intensity assessed using the visual analogue scale (VAS), white blood cell (WBC) count, C-reactive protein (CRP), procalcitonin (PCT), swelling regression time, pain relief time, bacterial clearance, lesion-site subgroup responses, and safety. Percentage reduction in lesion swelling area was calculated at the participant level as (baseline swelling area minus day 7 swelling area) divided by baseline swelling area and multiplied by 100. VAS pain score decrease was calculated as baseline VAS score minus day 7 VAS score. Swelling regression time was defined as the time to >=80% reduction in swelling area compared with baseline. Pain relief time was defined as the time to reduction of local pain intensity to VAS <=3 sustained for at least 24 h.

#### 2.6.3. Bacteriological Assessment

Baseline and post-treatment specimens were collected aseptically from lesion exudate, wound surface swabs, or aspirated material when clinically appropriate and when no drainage indication was present. Bacterial culture and identification were performed using standard laboratory procedures and an automated microbial identification system. Methicillin resistance in *Staphylococcus aureus* was determined by cefoxitin disk diffusion according to Clinical and Laboratory Standards Institute (CLSI) M100 [16]. Bacterial clearance was defined as the absence of baseline pathogens in available post-treatment culture specimens. Participants without baseline pathogen-positive cultures were not included in pathogen-specific clearance denominators. Because not all participants had microbiologically evaluable specimens and pathogen-specific sample sizes were limited, microbiological outcomes were regarded as exploratory.

#### 2.6.4. Safety Assessment

Safety was evaluated throughout the 7-day treatment course through daily clinical assessment, adverse-event monitoring, and clinically indicated laboratory evaluation when necessary. Monitored safety outcomes included local irritation, itching, burning sensation, erythema aggravation, rash, allergy, gastrointestinal symptoms, infusion reactions, and laboratory abnormalities identified during routine clinical care. Skin irritation was graded according to Chinese technical guidance for irritation testing [17]. Serious adverse events were defined as death, life-threatening events, hospitalization prolongation, permanent disability, or events requiring urgent intervention. All adverse events were recorded regardless of presumed causality. No predefined stopping rules were triggered during the study period.

### 2.7. Sample Size Calculation

Sample size was calculated using G*Power version 3.1 (Heinrich Heine University Düsseldorf, Germany) based on the primary outcome of primary clinical response rate. According to preliminary clinical observations and the expected clinically meaningful between-group differences, the anticipated response rates were estimated as 90% for the combined group, 75% for the *T. mongolicum* group, and 65% for the ceftriaxone group. Sample size estimation was performed using the chi-square test for comparison of proportions among three independent groups, with a significance level (α) of 0.05, statistical power (1 − β) of 0.80, and an allocatio ratio of 1:1:1. The calculated minimum sample size was 52 patients per group. Assuming a potential dropout rate of approximately 15%, 60 patients were enrolled in each group, resulting in a total planned sample size of 180 patients. Ultimately, 175 patients completed the study, whereas all 180 randomized patients were included in the intention-to-treat (ITT) analysis.

### 2.8. Statistical Analysis

All statistical analyses reported in this manuscript were performed after trial registration. The analysis followed the intention-to-treat (ITT) principle. All randomized participants were included in the primary analysis according to their randomized allocation, regardless of treatment adherence, withdrawal, or receipt of rescue treatment. The primary estimand was interpreted using a treatment-policy approach: clinical deterioration requiring rescue treatment or protocol discontinuation for clinical reasons was classified as non-response. Missing post-treatment continuous data were handled using the last observation carried forward (LOCF) method for the primary analysis of continuous secondary outcomes; the limitations of this approach are acknowledged in the Discussion. Given the small amount of incomplete follow-up, post hoc sensitivity analyses for the primary binary outcome were performed using (1) complete cases only and (2) a conservative principal-comparison scenario in which non-completers in the combined group were classified as non-responders and the non-completer in the ceftriaxone group was classified as a responder.

Continuous variables are presented as mean ± standard deviation (SD) and were compared using one-way analysis of variance (ANOVA) when assumptions were satisfied. Categorical variables are presented as n (%) and were compared using Pearson’s chi-square test or Fisher’s exact test when appropriate. Post hoc pairwise comparisons for continuous outcomes were derived from the ANOVA pooled error term, and pairwise comparisons for categorical outcomes used Pearson’s chi-square tests; pairwise comparisons were interpreted cautiously using a Bonferroni-adjusted threshold of P < 0.0167. The principal clinically relevant pairwise comparison was defined as the combined group versus the ceftriaxone group, because the study was designed to evaluate the adjunctive value of topical *T. mongolicum* wet dressing when added to systemic ceftriaxone. Comparisons involving the *T. mongolicum* monotherapy arm, lesion-site subgroup analyses, and microbiological subgroup analyses were considered supportive or exploratory, were not adjusted for multiplicity, and should be interpreted as hypothesis-generating only. Statistical analyses were performed using IBM SPSS Statistics version 26.0 (IBM Corp, Armonk, NY, USA). Statistical reporting followed ICH E9 and E9(R1) principles [18,19]. The sample size was primarily designed to support the overall three-group comparison and was not specifically powered for multiplicity-adjusted pairwise comparisons or exploratory subgroup analyses.

## 3. Results

### 3.1. Study Population and Baseline Characteristics

A total of 207 patients were screened for eligibility. Among them, 27 were excluded (20 did not meet the inclusion criteria and 7 declined participation). The remaining 180 patients were randomly assigned equally into the combined group, *T. mongolicum* group, and ceftriaxone group (n = 60 per group) (Fig 1).

**Fig 1.**
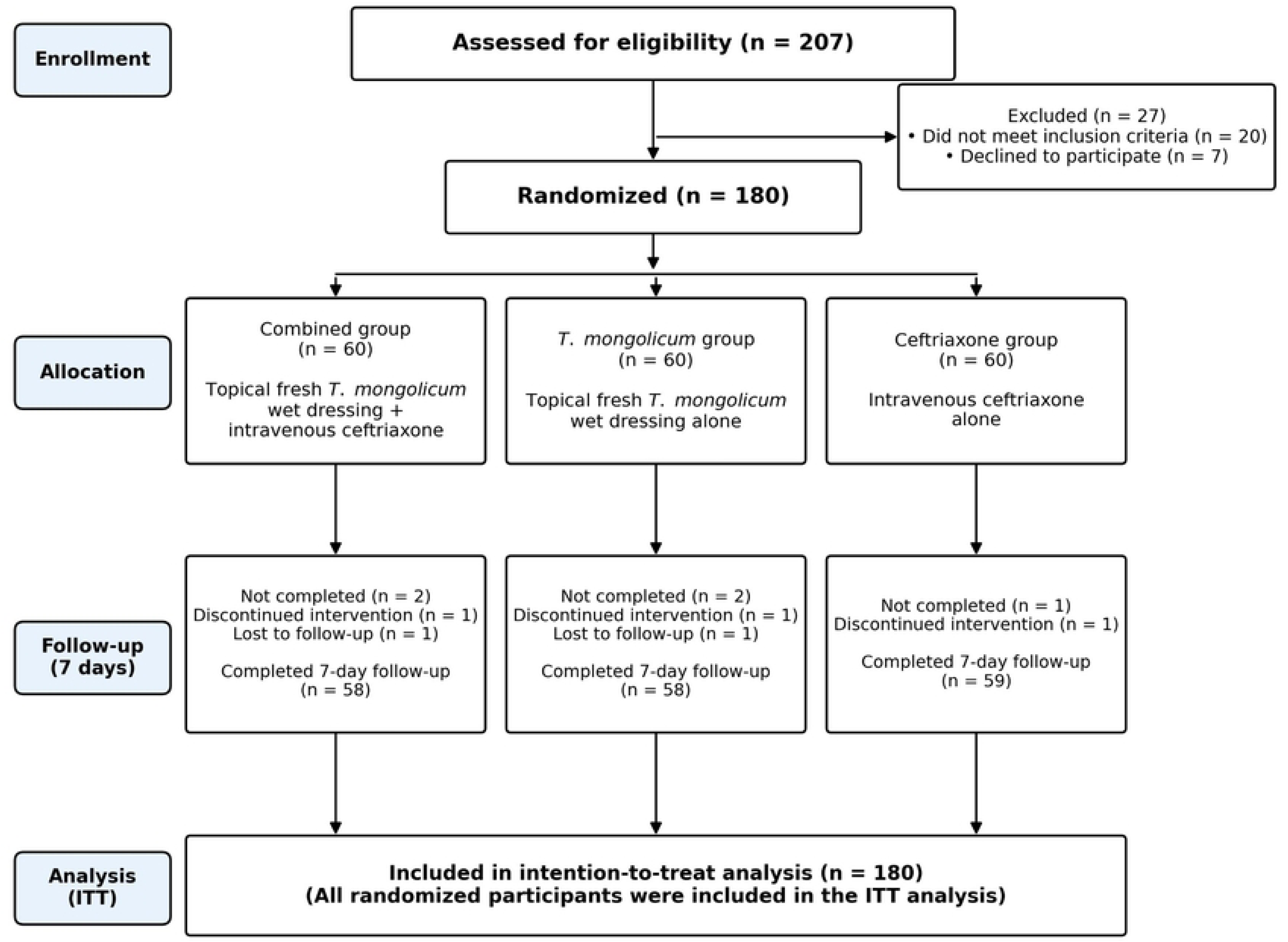
CONSORT flow diagram of participant enrollment, allocation, follow-up, and analysis. Note,: Participant disposition reasons are mutually exclusive and defined in the 54 Dataset (variable: primary_disposition_reason). ·Abbreviations: ITT. intention-to-treat; *T. mongolicum, Taraxacum mongolicum*.

Fig 1. CONSORT flow diagram of participant enrollment, allocation, follow-up, and analysis. Baseline demographic characteristics, disease duration, lesion-site distribution, baseline swelling area, and baseline VAS pain score were comparable among the three groups, with no statistically significant differences observed at enrollment (Table 1).

**Table 1.** Baseline characteristics of the three groups (mean ± SD or n [%]).

| Characteristic | Combined group<br>(n=60) | *T. mongolicum* group<br>(n=60) | Ceftriaxone<br>group (n=60) | F/$\chi^2$ | P value |
| --- | --- | --- | --- | --- | --- |
| Sex (male/female) | 32/28 | 33/27 | 31/29 | 0.13 | 0.935 |
| Age (years) | 45.76 ± 12.89 | 46.12 ± 13.05 | 45.43 ± 12.76 | 0.04 | 0.958 |
| Course of disease (days) | 9.87 ± 3.01 | 9.66 ± 3.15 | 10.01 ± 2.87 | 0.21 | 0.815 |
| Baseline VAS pain score | 6.72 ± 1.01 | 6.66 ± 1.04 | 6.61 ± 1.00 | 0.18 | 0.839 |
| Lesion site distribution [n (%)] |  |  |  | 0.27 | 1.000 |
| Head/face/neck | 16 (26.67) | 15 (25.00) | 16 (26.67) |  |  |
| Trunk | 14 (23.33) | 16 (26.67) | 15 (25.00) |  |  |
| Extremities | 15 (25.00) | 14 (23.33) | 15 (25.00) |  |  |
| Perineum/buttocks region | 15 (25.00) | 15 (25.00) | 14 (23.33) |  |  |
| Baseline swelling area (cm<sup>2</sup>) | 45.26 ± 18.35 | 44.89 ± 17.92 | 45.11 ± 18.07 | 0.01 | 0.994 |

Notes: Continuous variables are presented as mean ± standard deviation (SD), and categorical variables as n (%). Baseline analyses were performed on the intention-to-treat (ITT) population. Detailed lesion-site subgroup baseline data are provided in Supplementary Table S1.

### 3.2. Primary Outcome

After 7 days of treatment, the primary clinical response rate in the ITT population was 91.67% (55/60) in the combined group, 76.67% (46/60) in the *T. mongolicum* group, and 68.33% (41/60) in the ceftriaxone group. The overall intergroup difference was statistically significant (χ² = 10.07, P = 0.006). In the principal clinically relevant comparison, the combined group was associated with a higher response rate than the ceftriaxone group, with a risk difference of 23.3 percentage points (95% CI, 9.6 to 37.0), risk ratio of 1.34 (95% CI, 1.11 to 1.62), and odds ratio of 5.10 (95% CI, 1.76 to 14.79). For the primary clinical response outcome, the comparisons of combined therapy versus *T. mongolicum* monotherapy (P = 0.024) and *T. mongolicum* monotherapy versus ceftriaxone monotherapy (P = 0.307) were considered supportive or exploratory and did not meet the Bonferroni-adjusted significance threshold of P < 0.0167 (Table 2).

**Table 2.**
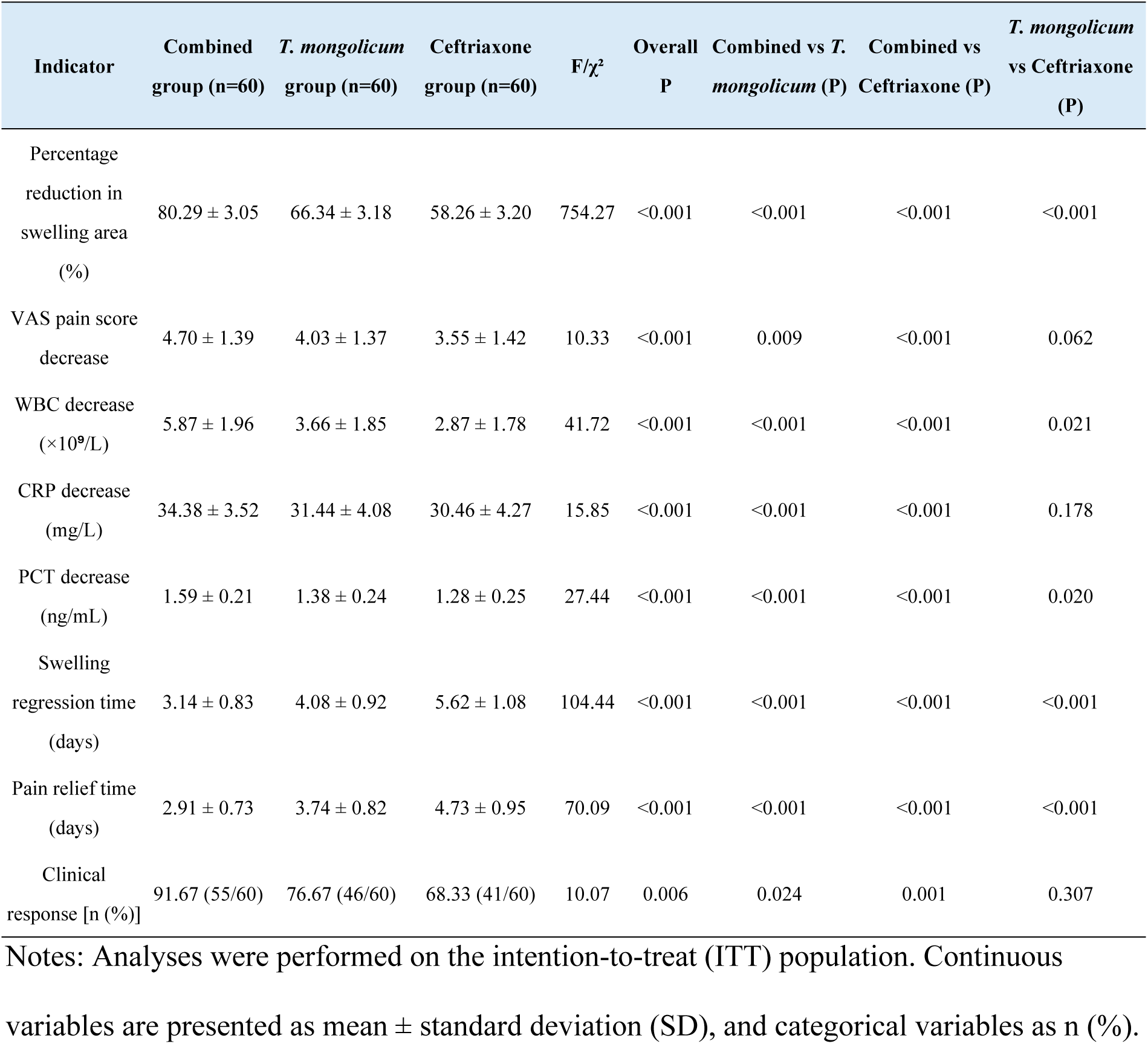
Primary and key secondary outcomes across the three treatment groups, with pairwise comparisons (mean ± SD or n [%]).

Post hoc sensitivity analyses supported the robustness of the principal comparison. In the complete-case analysis, clinical response rates were 94.83% (55/58), 79.31% (46/58), and 69.49% (41/59) in the combined, *T. mongolicum*, and ceftriaxone groups, respectively (overall P = 0.002; combined versus ceftriaxone, P < 0.001). In a conservative principal-comparison scenario that classified non-completers in the combined group as non-responders and the non-completer in the ceftriaxone group as a responder, the combined group remained associated with a higher clinical response rate than ceftriaxone alone (55/60 [91.67%] versus 42/60 [70.00%]; P = 0.003). Detailed sensitivity analyses are provided in Supplementary Table S2.

### 3.3. Secondary Clinical, Inflammatory, and Recovery Outcomes

Secondary clinical, inflammatory, and recovery outcomes were directionally consistent with the primary analysis. The combined group showed greater percentage reduction in lesion swelling area and greater VAS pain score decrease than both monotherapy groups. Reductions in WBC, CRP, and PCT were also more pronounced in the combined group. Compared with ceftriaxone monotherapy, *T. mongolicum* monotherapy showed a greater percentage reduction in swelling area and shorter swelling regression and pain relief times; the VAS pain score decrease was numerically greater but did not meet the Bonferroni-adjusted significance threshold. These secondary outcomes were considered supportive of the primary analysis. Comparisons involving the topical monotherapy arm should be interpreted cautiously because they were supportive or exploratory rather than the prespecified principal clinical comparison (Table 2; Supplementary Tables S3 and S4).

Percentage reduction in swelling area was calculated at the participant level as (baseline swelling area minus day 7 swelling area) divided by baseline swelling area and multiplied by 100. VAS pain score decrease was calculated as baseline VAS score minus day 7 VAS score. Post hoc pairwise comparisons were interpreted using a Bonferroni-adjusted significance threshold of P < 0.0167. For the primary clinical response, the principal comparison of combined therapy versus ceftriaxone monotherapy additionally showed a risk difference of 23.3 percentage points (95% CI, 9.6 to 37.0), risk ratio of 1.34 (95% CI, 1.11 to 1.62), and odds ratio of 5.10 (95% CI, 1.76 to 14.79).

### 3.4. Microbiological Outcomes

Baseline bacteriological specimens were collected whenever clinically feasible before treatment initiation. *Staphylococcus aureus* was the most frequently isolated pathogen, identified in 107 patients overall, including 25 MRSA isolates. *Escherichia coli* was identified in 33 patients. Baseline pathogen distribution did not differ significantly among the three groups (all P > 0.05). In the ITT summary, total bacterial clearance differed among groups and was numerically highest in the combined group [81.67% (49/60)], followed by the *T. mongolicum* group [70.00% (42/60)] and the ceftriaxone group [58.33% (35/60)] (χ² = 7.78, P = 0.020). Because microbiological outcomes were exploratory, not adjusted for multiplicity, and limited by incomplete culture-evaluable denominators, these results should be interpreted as supportive rather than confirmatory. Pathogen-specific clearance rates showed numerically favorable trends in the combined group, but these analyses were exploratory because of limited culture-evaluable subgroup sizes (Table 3).

**Table 3.** Baseline pathogen distribution and post-treatment bacteriological clearance.

| Variable | Combined group | *T. mongolicum* group | Ceftriaxone group | $\chi^2$ | P value |
| --- | --- | --- | --- | --- | --- |
| *S. aureus* at baseline | 37 (61.67) | 35 (58.33) | 35 (58.33) | 0.18 | 0.912 |
| MRSA among *S. aureus* isolates | 8 (13.33) | 10 (16.67) | 7 (11.67) | 0.65 | 0.722 |
| *E. coli* at baseline | 10 (16.67) | 11 (18.33) | 12 (20.00) | 0.22 | 0.895 |
| Other pathogens | 10 (16.67) | 8 (13.33) | 9 (15.00) | 0.26 | 0.877 |
| Multiple pathogens | 5 (8.33) | 4 (6.67) | 3 (5.00) | 0.54 | 0.765 |
| Total bacterial clearance | 49/60 (81.67) | 42/60 (70.00) | 35/60 (58.33) | 7.78 | 0.020 |
| *S. aureus* clearance | 31/37 (83.78) | 27/35 (77.14) | 23/35 (65.71) | 3.25 | 0.197 |
| MRSA clearance | 6/8 (75.00) | 6/10 (60.00) | 3/7 (42.86) | 1.61 | 0.448 |
| *E. coli* clearance | 8/10 (80.00) | 8/11 (72.73) | 7/12 (58.33) | 1.28 | 0.526 |

Notes: MRSA was counted as a methicillin-resistant subset of *S. aureus*. Multiple-pathogen infections were not mutually exclusive with individual pathogen categories. Other pathogens included coagulase-negative staphylococci (n = 12), *Streptococcus pyogenes* (n = 9), *Proteus mirabilis* (n = 4), and *Klebsiella pneumoniae* (n = 2). Microbiological subgroup analyses were exploratory because of limited pathogen-specific sample sizes.

### 3.5. Exploratory Subgroup Analysis by Lesion Site

#### 3.5.1 Primary Clinical Response by Lesion Site

Exploratory lesion site-stratified analysis suggested possible variation in primary clinical response across anatomical regions (Table 4). In the combined group, primary clinical response was numerically higher in extremity and head/face/neck lesions than in trunk or perineal/gluteal lesions. A similar but less pronounced numerical pattern was observed in the *T. mongolicum* group, whereas no clear lesion site-related pattern was evident in the ceftriaxone group. These findings should be considered hypothesis-generating because the subgroup analyses were exploratory, had limited sample sizes, and were not adjusted for multiplicity.

**Table 4.**
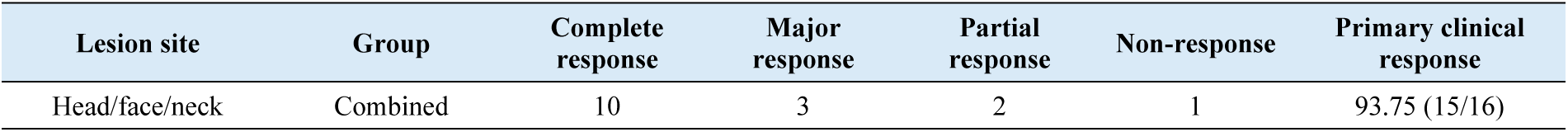
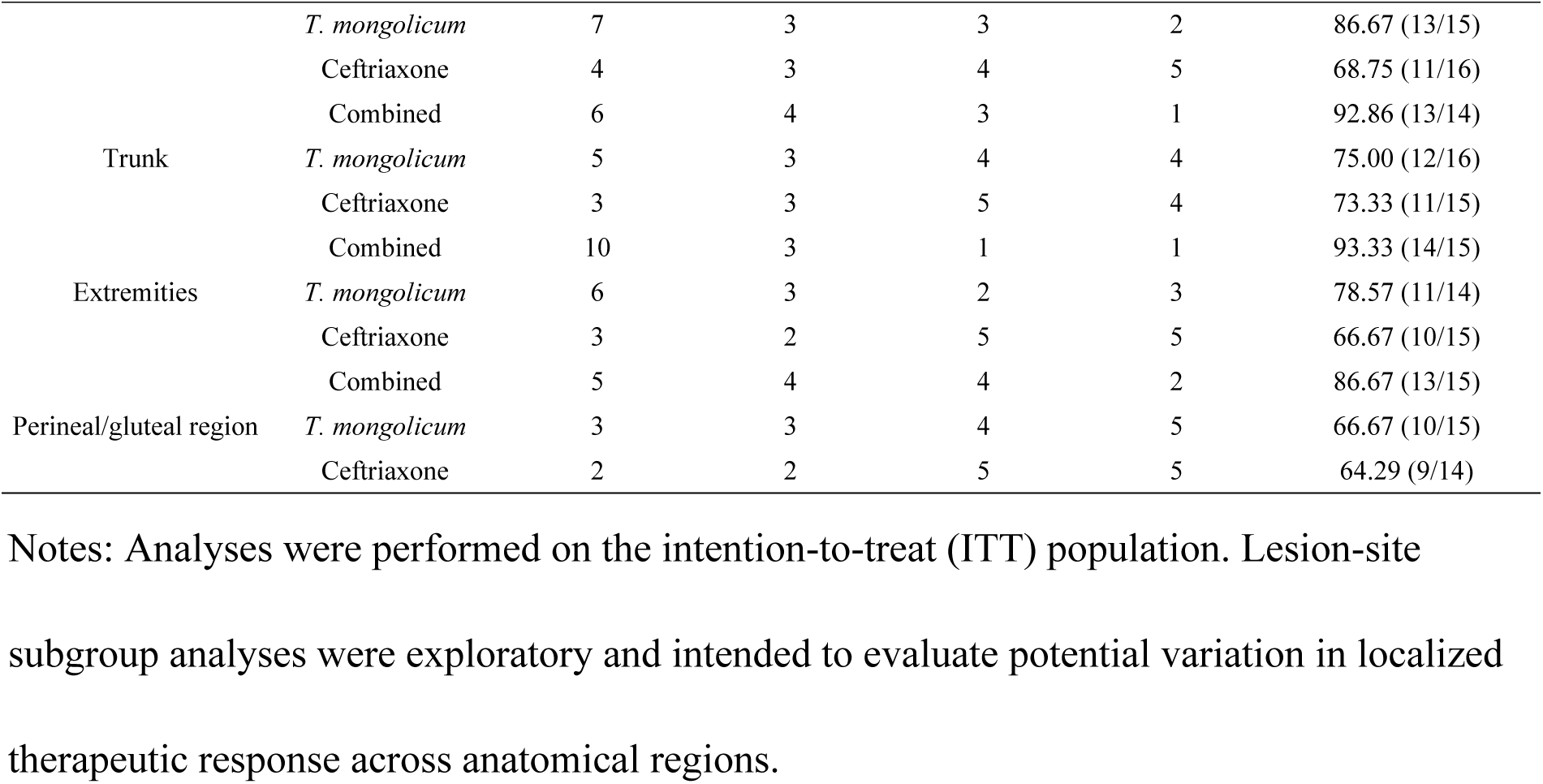
Comparison of primary clinical response among lesion-site subgroups [n (%)].

#### 3.5.2 Swelling Area Percentage Reduction, Pain Intensity, and Symptom Recovery

Exploratory lesion site-stratified analyses suggested that anatomical variation may have been more apparent for recovery-time outcomes than for fixed day-7 percentage reduction in local swelling area or VAS pain score decrease. In the combined and *T. mongolicum* groups, swelling regression time and pain relief time were numerically shorter for head/face/neck and extremity lesions than for trunk or perineal/gluteal lesions. Differences in day-7 percentage reduction in swelling area and VAS pain score decrease were less consistent across lesion sites. These findings should be interpreted cautiously because the subgroup analyses were exploratory, underpowered, and not adjusted for multiplicity (Table 5).

**Table 5.**
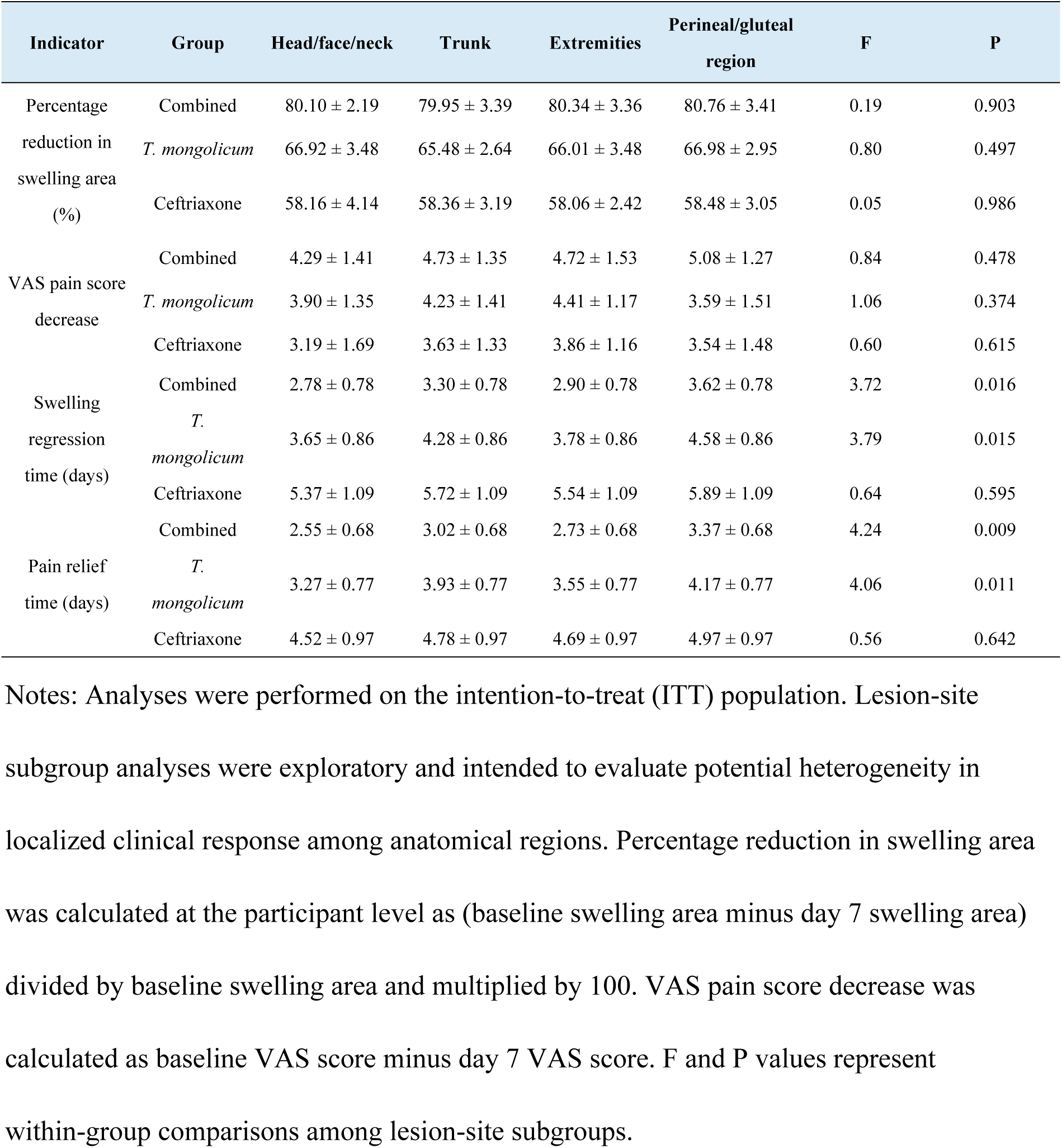
Comparison of percentage reduction in swelling area, VAS pain score decrease, and symptom recovery among lesion-site subgroups (mean ± SD).

#### 3.5.3. Inflammatory Indicator Reduction by Lesion Site

Exploratory lesion site-stratified analyses of inflammatory indicators demonstrated less consistent heterogeneity than that observed for symptom recovery indicators. In the combined group, modest lesion site-related numerical differences were observed for some inflammatory biomarkers, whereas biomarker subgroup patterns in the *T. mongolicum* and ceftriaxone groups were less apparent. Because multiple subgroup comparisons were performed with small sample sizes, these findings should not be used for definitive subgroup inference (Table 6).

**Table 6.** Exploratory lesion site-specific inflammatory indicator reduction (mean ± SD).

| Indicator | Group | Head/face/neck | Trunk | Extremities | Perineal/gluteal region | F | P |
| --- | --- | --- | --- | --- | --- | --- | --- |
| WBC decrease<br>($\times 10^9/L$) | Combined | 6.25 ± 1.99 | 5.68 ± 1.99 | 6.02 ± 1.99 | 5.50 ± 1.99 | 0.44 | 0.726 |
| | *T. mongolicum* | 3.85 ± 1.89 | 3.52 ± 1.89 | 3.73 ± 1.89 | 3.57 ± 1.89 | 0.10 | 0.961 |
| | Ceftriaxone | 2.94 ± 1.82 | 2.80 ± 1.82 | 2.99 ± 1.82 | 2.75 ± 1.82 | 0.06 | 0.982 |
| CRP decrease<br>(mg/L) | Combined | 36.03 ± 3.35 | 33.63 ± 3.35 | 35.03 ± 3.35 | 32.68 ± 3.35 | 3.00 | 0.038 |
| | *T. mongolicum* | 32.33 ± 4.13 | 31.11 ± 4.13 | 31.79 ± 4.13 | 30.56 ± 4.13 | 0.53 | 0.666 |
| | Ceftriaxone | 30.81 ± 4.37 | 30.28 ± 4.37 | 30.66 ± 4.37 | 30.02 ± 4.37 | 0.10 | 0.959 |
| PCT decrease<br>(ng/mL) | Combined | 1.66 ± 0.21 | 1.55 ± 0.21 | 1.62 ± 0.21 | 1.52 ± 0.21 | 1.45 | 0.238 |
| | *T. mongolicum* | 1.43 ± 0.24 | 1.37 ± 0.24 | 1.41 ± 0.24 | 1.32 ± 0.24 | 0.60 | 0.619 |
| | Ceftriaxone | 1.32 ± 0.25 | 1.27 ± 0.25 | 1.30 ± 0.25 | 1.24 ± 0.25 | 0.28 | 0.839 |

Notes: Analyses were performed on the intention-to-treat (ITT) population. Biomarker subgroup analyses were exploratory and intended to assess potential heterogeneity in localized inflammatory response among anatomical regions. F and P values represent within-group comparisons among lesion-site subgroups.

### 3.6. Safety

No serious adverse events or rescue-treatment-related safety events were reported during the 7-day treatment period. No adverse event required treatment discontinuation, rescue treatment, or emergency intervention. Mild self-limited local skin symptoms and infusion-site discomfort were noted during routine clinical monitoring, but non-serious adverse events were not collected as prespecified participant-level quantitative endpoints; therefore, safety findings should be interpreted as short-term descriptive safety data.

## 4. Discussion

This single-center randomized controlled trial found that standardized topical fresh *T. mongolicum* wet dressing, when used as an adjunct to systemic ceftriaxone, was associated with improved short-term clinical outcomes in selected clinically stable adults with localized SSTIs. Compared with ceftriaxone monotherapy, the combined regimen was associated with a higher day-7 clinical response rate, greater reduction in swelling area, faster symptom relief, and greater reductions in inflammatory biomarkers during the 7-day treatment period. These findings suggest a potential adjunctive role for topical *T. mongolicum* wet dressing in a restricted clinical setting, but they should not be interpreted as evidence that the dressing can replace antibiotics, drainage, or urgent clinical care when these are indicated.

The three-arm design was intended primarily to evaluate the adjunctive value of topical *T. mongolicum* when added to ceftriaxone. The topical monotherapy arm provides supportive information about the possible local contribution of the wet dressing, but it was restricted to clinically stable participants without systemic infection, deep tissue involvement, rapidly progressive disease, or drainage indications. Accordingly, comparisons involving the topical monotherapy arm should be interpreted cautiously and should not be generalized to SSTI populations requiring systemic antibacterial therapy or surgical management.

The observed differences with the combined regimen may reflect complementary effects of systemic antibacterial treatment and localized topical intervention. While ceftriaxone provides systemic antimicrobial coverage against common SSTI pathogens [1], topical *T. mongolicum* may contribute to local symptom improvement through anti-inflammatory or microenvironment-modulating effects suggested by preclinical studies [7–9,20,21]. However, mechanistic interpretations remain speculative because this trial did not include pharmacodynamic measurements, skin penetration studies, cytokine profiling, or a placebo wet-dressing control. Therefore, the clinical findings should be interpreted as evidence of association rather than proof of a specific mechanism.

Microbiological findings suggested a numerical trend toward higher bacterial clearance with combination therapy. The combined regimen had numerically higher clearance rates for common SSTI pathogens, including *Staphylococcus aureus* and MRSA. However, pathogen-specific analyses were limited by sample size and by the fact that not all participants had microbiologically evaluable specimens. These microbiological findings are therefore exploratory and require confirmation in studies with complete microbiological sampling and prespecified pathogen-specific endpoints.

Exploratory analyses suggested possible heterogeneity in treatment response across anatomical lesion sites. Lesions in head/face/neck and extremity regions appeared to have numerically shorter symptom recovery times in the topical-treatment groups than lesions in trunk or perineal/gluteal regions. This observation may be biologically plausible because regional differences in skin permeability, microcirculation, tissue thickness, and local mechanical stress can influence topical exposure and symptom resolution [22,23]. Nevertheless, these subgroup analyses were not powered for definitive inference and were not adjusted for multiplicity; they should be considered hypothesis-generating only.

Several strengths of this study should be acknowledged. The randomized controlled design enabled direct comparison between adjunctive combination therapy and ceftriaxone monotherapy. Randomization was stratified by lesion site, and outcome assessments were performed by blinded independent assessors. The study also used prespecified intervention regimens, quality-controlled botanical preparation, intention-to-treat analysis, CONSORT documentation, an approved protocol, and a deidentified participant-level dataset that allows independent reproduction of the main findings.

Several limitations should be considered. First, this was a single-center study with a short 7-day follow-up period, which limits generalizability and does not allow assessment of recurrence, relapse, or longer-term safety. Second, the trial did not include a placebo or normal-saline wet-dressing control group; therefore, it cannot distinguish specific pharmacological effects of *T. mongolicum* from nonspecific effects related to moist dressing, local cooling, occlusion, or additional clinical attention. Third, participants and treating physicians were not blinded because of the different routes of administration, introducing potential performance bias. Although outcome assessors and statisticians were blinded and objective secondary outcomes were directionally consistent with the primary outcome, residual bias cannot be excluded.

Fourth, the primary clinical response outcome incorporated global clinical improvement criteria and therefore included subjective clinical judgment, despite prespecified definitions and blinded assessment. Fifth, public trial registration occurred after participant enrollment had begun; although the protocol and statistical analysis plan were finalized before enrollment and were not changed after registration, future trials should be registered prospectively before participant enrollment. Sixth, LOCF was used for some missing post-treatment continuous data, and more robust missing-data methods should be incorporated into future trials. Seventh, non-serious adverse events were monitored clinically but were not collected as prespecified participant-level quantitative endpoints, limiting detailed safety comparisons. Finally, lesion-site and pathogen-specific analyses were exploratory, underpowered, and not adjusted for multiplicity.

Future studies should incorporate multicenter designs, larger sample sizes, longer follow-up periods, placebo-controlled topical comparator arms, complete microbiological sampling, and prespecified sensitivity analyses. Mechanistic studies are also needed to clarify whether any observed clinical benefit is attributable to specific botanical pharmacological effects, nonspecific wet-dressing effects, or a combination of both.

## 5. Conclusions

In this single-center randomized trial, adjunctive topical fresh *T. mongolicum* wet dressing combined with ceftriaxone was associated with improved short-term clinical outcomes compared with ceftriaxone alone in selected clinically stable adults with localized SSTIs. These findings suggest a potential adjunctive role for standardized topical botanical therapy; however, they should be interpreted cautiously given the single-center design, short follow-up period, lack of placebo control, partial blinding, and exploratory nature of several secondary analyses. The dressing should not be considered a replacement for antibiotics, drainage procedures, or urgent clinical care when indicated. Multicenter, placebo-controlled studies with larger sample sizes, longer follow-up, complete microbiological denominators, and mechanistic investigations are warranted to validate these findings and clarify the role of *T. mongolicum* in SSTI management.

Supporting information files are listed after the References. S3 File contains supplementary Tables S1-S5, including lesion-site subgroup baseline characteristics, baseline and post-treatment inflammatory indicators, swelling and pain recovery indicators, batch-level botanical quality-control records, and primary outcome sensitivity analyses.

Author contributions: Conceptualization: Shengcheng Ma, Han Yang. Methodology: Shengcheng Ma, Han Yang. Investigation: Yi Wang, Xiaoqing Xian, Shengjun Nie. Data curation: Yi Wang, Xiaoqing Xian, Shengjun Nie. Formal analysis: Yi Wang, Xiaoqing Xian, Shengjun Nie. Visualization: Xiaoqing Xian, Shengjun Nie. Writing – original draft: Yi Wang. Writing – review & editing: Yi Wang, Xiaoqing Xian, Shengjun Nie, Shengcheng Ma, Han Yang. Supervision: Shengcheng Ma, Han Yang. Project administration: Shengcheng Ma, Han Yang. Funding acquisition: Shengcheng Ma, Han Yang.

## Ethics statement

The study was conducted in accordance with the Declaration of Helsinki and was approved by the Ethics Committee of the 943rd Hospital of the Joint Logistic Support Force of the Chinese People’s Liberation Army (protocol code PLA943EC24002; approval date: 8 November 2024). Written informed consent was obtained from all participants before enrollment.

## Data availability statement

All relevant data are within the article and its Supporting Information files. The deidentified participant-level minimal dataset underlying the reported findings, together with the data dictionary and codebook, is provided as S4 Dataset. Aggregate data supporting the main and supplementary tables are included in the article and S3 File. No separate statistical code file was generated; the analysis methods are described in Section 2.8.

## Acknowledgments

Not applicable.

## Supporting information

S1 Checklist. CONSORT checklist.

S2 Protocol. Trial protocol approved by the ethics committee.

S3 File. Supplementary tables and botanical quality-control data. This file contains Tables S1-S5, including lesion-site subgroup baseline characteristics, baseline and post-treatment inflammatory indicators, swelling and pain recovery indicators, batch-level quality-control records for fresh *T. mongolicum* used during the trial, and primary outcome sensitivity analyses.

S4 Dataset. Deidentified minimal participant-level dataset and data dictionary.

